# The definition and risks of Cytokine Release Syndrome-Like in 11 COVID-19-Infected Pneumonia critically ill patients: Disease Characteristics and Retrospective Analysis

**DOI:** 10.1101/2020.02.26.20026989

**Authors:** Wang Wenjun, Liu Xiaoqing, Wu Sipei, Lie Puyi, Huang Liyan, Li Yimin, Cheng Linling, Chen Sibei, Nong Lingbo, Lin Yongping, He Jianxing

**Affiliations:** Guangzhou Respiratory Health Research Institute, The First Affiliated Hospital of Guangzhou Medical University, State Key Laboratory of respiratory diseases, Guangdong, China. 510120; Department of Critical Care Medicine,, the First Affiliated Hospital of Guangzhou Medical University, Guangdong, China.510120; Guangdong Provincial People’s Hospital, Guangdong Academy of Medical Sciences, Guangdong, China. 510080; Guangzhou Eighth People’s Hospital, Guangdong, China; Department of laboratory, the First Affiliated Hospital of Guangzhou Medical University, Guangdong, China. 510120

**Keywords:** COVID-19, Cytokine Release Syndrome-Like, IL-6, Immunophenotype, Severe/critical pneumonia

## Abstract

**IMPORTANCE:** COVID-19-infected pneumonia patients with severe immune abnormalities and risk of cytokine release syndrome. The definition, prevention, and treatment of COVID-19-infected pneumonia in critically ill patients with cytokine release syndrome symptoms is an important problem.

**OBJECTIVE:** To define the cytokine release syndrome-like (CRSL) in COVID-19-infected pneumonia in critically ill patients and study the risk factors and therapeutic strategies.

**DESIGN, SETTING, AND PARTICIPANTS:** This is a retrospective, single center case study of 11 COVID-19-infected pneumonia patients with acute respiratory distress syndrome (ARDS) from The First Affiliated Hospital of Guangzhou Medical University in China from January 26, 2020 to February 18, 2020. The follow-up termination date was February 19, 2020.

**EXPOSERES:** Eleven COVID-19-infected pneumonia patients with ARDS in the ICU. Some of these patients also had cytokine release syndrome-like (CRSL). Immunologic detection, clinical characteristics, and clinical treatment analysis were carried out to define the CRSL in these COVID-19-infected pneumonia patients.

**MAIN OUTCOMES AND MEASURES:** Clinical, radiological, immunology (including immune cell subsets and cytokines analysis), laboratory, and clinical treatment data were collected and analyzed. The critically ill patients with CRSL were defined. Prevention and control strategies were studied.

**RESULTS:** Of 11 critically ill patients in the ICU, the median age was 58 (Inter-Quartile Range{IQR}, 49–72; range, 26–72 years), and 10 (83.3%) were males. Ten (83.3%) patients had extensive pulmonary inflammation and ARDS (the median time from the first symptom to ARDS was 10.0 d), fever, and hypoxia; four (28.6%) patients experienced shock. The lymphocyte subpopulations including CD3 (CD3 + CD45+), CD4 (CD3 + CD4+), CD8 (CD3 + CD8+), NK (CD3-CD16 + CD56 +), Tregs (CD4 + CD25 + CD127 low), B lymphocyte (CD3-CD19 +) cells; and cytokines including IL-2, IL-4, IL-6, IL-10, TNF-α, IFN-γ were detected at different time points. All of the patients had a decrease of CD3 (IQR,169–335; range, 50–635 cells/μL), CD4 (IQR,101–303; range, 27–350 cells/μL), CD8 (IQR, 33–141; range, 21–277 cells/μL); ten (90.9%) patients have a decrease in NK immune cells (IQR,8–72; range, 5–170 cells/μL); both of Tregs (IQR,3.3-7.8;rang,2.3-9.4%) and B immune cells (IQR,61-146; rang,44-222 cells/μL)were decreased in two (18.2%) patients. And nine patients were increase in CD4 / CD8 (IQR,3.3–7.8%; range, 2.3–9.7%). All patients had a significant increase of IL-6 (IQR,14.26-92.2; range, 4.58–1182.91ng/L). Eight (72.7%) patients were determined to have CRSL characteristics, including pulmonary inflammation, fever, a decrease of CD4, CD8, and NK cells; an increase of CD4/CD8, a significant increase of IL-6, and the dysfunction of non-pulmonary organs. The numbers of CD4, CD8, and NK cells and the level of IL-6 in peripheral blood were correlated with the area of pulmonary inflammation in CT images *(P<0.05)*. Mechanical-ventilation used to increase blood oxygen concentration could increased the numbers of CD4 (after Vs before ventilation=259±53 VS 507±101; P=0.013, and CD8 (after Vs before ventilation=193±38 VS 279±63; P=0.048), while decreasing the level of IL-6 (after Vs before ventilation=223.2± 89.9 VS 26.8±10; P=0.041). The increased of IL-6 was occurred earlier than the decrease of CD4·, CD8 in the patients with rapidly worsened after ICU.

**CONCLUSIONS AND RELEVANCE:** In this retrospective analysis of 11 critically ill pneumonia patients infected with COVID-19, we defined and identified eight patients (72.7%) with cytokine release syndrome-like (CRSL). We found that a large area of lung injury (≥50%) with an decrease of CD4, CD8 (Lower than 50% minimum normal range) and increase of IL-6 in peripheral blood was the highest risk factor of CRSL. IL-6 was a early indicators of CRSL in COVID-19-infected pneumonia. We also found that reduce injury to the lung is a useful method to prevent and improve COVID-19-infected pneumonia-related CRSL in critically ill pneumonia patients.

---

In December 2019, a Novel Coronavirus which was named as the severe acute respiratory syndrome coronavirus 2 (COVID-19) by the International Committee on Taxonomy of Viruses occurred in Wuhan, Hubei province, China [1-3]. The disease spread rapidly to all provinces of China outside of Wuhan[4,5]. By February 17, 2020, there were 70,636 confirmed cases and 1,772 deaths in China [6]. Some of the COVID-19-infected critically ill pneumonia patients had serious symptoms with no specific drug treatment indicated. Severe inflammatory reaction and respiratory distress syndrome can lead to rapid progression of the disease and cause death. A guideline “Diagnosis and treatment of novel coronavirus pneumonia (trial version fifth)” was first published on February 3 in China [7]. It suggested that immune factors were needed to examine in the therapy of COVID-19-infected pneumonia patients. The release of inflammatory immune cytokines could increase the inflammatory of the lung and risk the occurrence of respiratory distress syndrome (ARDS). ARDS could lead to hypoxia and lung injury, causing the release of inflammatory factors by multiple mechanisms [8-10]. So, an excess release of inflammatory factors may promote ARDS in COVID-19-infected pneumonia patients.

Critically ill COVID-19-infected pneumonia patients suffer from fever, pulmonary inflammation, respiratory distress, and other clinical symptoms. In published data, the proportion of ICU patients is 26.1% [11]. The average age of patients in the ICU is higher than that in non-ICU patients[11]. In addition, the incidence of acute heart injury and respiratory distress in ICU patients is much higher than that in the general population[12]. Serious pulmonary inflammation is a common symptom in critically ill COVID-19-infected pneumonia patients[13]. The large area of infection and inflammatory reaction will cause many immunologic problems, such as cytokine release syndrome (CRS), which can rapidly lead to deterioration and death[14]. In this study, after analysis of the clinical diagnosis and immunological characteristics of 11 COVID-19-infected patients with ARDS, we diagnosed eight (8/11, 72.7%) critically ill COVID-19-infected pneumonia patients to have cytokine release syndrome-like (CRSL) features. Therefore, we defined this phenomenon as COVID-19 infection-related CRSL. Our results found that IL-6 was a early indicators of CRSL in COVID-19-infected pneumonia, and suggestion that improving ventilation and controlling the area of pulmonary inflammation may be an effective method to treat the COVID-19-infected pneumonia patients with CRSL and ARDS. Our research provides experimental support and experiences useful for the treatment of critically ill COVID-19-infected pneumonia patients.

## Methods

### Study design and participants

This was a retrospective single-center study. The patients enrolled in this study were confirmed to have severe or critical COVID-19-infected pneumonia patients. The inclusion criteria were “Pneumonia diagnosis and treatment plan for new coronavirus infection (trial version 5)” issued by the National Health Commission on February 5, 2020 (hereinafter referred to as “Diagnosis and treatment plan version 5”) [7]. Enrollment began on January 26, 2020. We obtained verbal consent from each patient. According to the monitoring results (the date of transfer out of the ICU), the final follow-up date was February 18, 2020 for all COVID-19-infected patients participating in this study. This series of cases was approved by the ethics committee of the First Affiliated Hospital of Guangzhou Medical University.

### Data collection

The team of the First Affiliated Hospital of Guangzhou Medical University analysis of the patient’s medical records included:

a. Basic information of the patient: name, gender, age, hospitalization number, diagnosis, date of admission, condition, laboratory examination results, chest CT scan, nursing care, and treatments (i.e. antiviral treatment, corticosteroid treatment, and respiratory support. The records were taken by a team of trained doctors, and the data were obtained from electronic medical records.
b. Data generated by bedside monitor: heart rate, respiration, invasive / non-invasive blood pressure, blood oxygen saturation, hemodynamic monitoring.
c. Medical order execution information: administration times for all medicines, drug name, dose, concentration, route, automatically generated in and out quantity. These data were automatically collected by the ICU clinical information system.

Acute respiratory distress syndrome (ARDS) and shock are based on the World Health Organization Interim guidelines for the novel coronavirus. Hypoxemia was defined as PaO_2_/ FiO_2_ below 300 mm Hg. (PaO_2_ means arterial oxygen tension; FiO_2_ means inhaled oxygen fraction). Acute renal injury was determined and classified according to the highest serum creatinine level or urination standard and classified according to the global results of kidney disease improvement. If the patient had clinical symptoms or signs of nosocomial pneumonia or bacteremia, it was diagnosed as a secondary infection and combined with positive culture of new pathogens from lower respiratory tract samples (including sputum, tracheal intubation suction fluid, bronchoalveolar lavage fluid, or blood samples ≥ 48 hours after admission). If the serum level of cardiac biomarkers (such as cardiac troponin I) was higher than the 99% reference upper limit, or the ECG and echocardiography showed new abnormalities, the patient was diagnosed with cardiac injury.

### RT-PCR Assay for COVID-19

RT-PCR was performed with a Bio-RadCFX96 Real-Time PCR System, and 20 μL reaction volumes contained 10□μL of RNA, 5□μL ORF1ab/N primer provided with a COVID-19 kit (HuiruiBio, Shanghai,China), 1 detection reaction power with one step RT-PCR Master Mix Reagents from the kit, and 5□μL RNase Free H_2_O (TiangenBio, Beijing, China). Thermal cycling was performed at 55°C for 15 minutes for reverse transcription, followed by 95°C for 5 minutes, 45 cycles of 95°C for 10 seconds, and then 55°C for 40 seconds. Fluorescence was recorded during the 55°C phase. The results of the RT-PCR were valued by threshold cycle (CT). When CT>39.2, the sample was negative; when CT<35, the sample was positive; when 35<CT<39.2, the sample was retested.

### Flow cytometry analysis

EDTA anticoagulation venous blood samples (2 mL) were collected from each of the 2019-nCoV patients. Peripheral blood mononuclear cells (PBMCs) were isolated using density-gradient Ficoll-Paque Plus (Amersham Biosciences, Little Chalfont, United Kingdom) centrifugation. Duplicate PBMCs (10^6^ cells/tube) were stained with CD3-FITC/CD56+16-PE/CD45-PerCP-CY5.5/CD4-PC7/CD19-APC/CD8-APC -CY7/CD25-APC/CD127-PE (Tongsheng Shidai Biote Co. LTD, Beijing, China) in the dark at room temperature for 30□minutes. The cells were then fixed and permeabilized using a permeabilization solution (BD Biosciences, New Jersey, USA). The mononuclear cells were gated using cell size and internal structure in a FACS Aria II (Novo Cyte D3000, Agilent Technologies Inc., Palo Alto, California, USA). The frequency of various subsets of T cells was determined using FlowJo software (v10; TreeStar, Ashland, OR, USA). The number of different subsets of T cells was calculated and expressed as the number of cells per L for each subject.

#### Cytokine detection

The plasma levels of IL-1β, IL-2, IL-4, IL-5, IL-6, IL-8, IL-10, IL-12P70, IL-17A, IFN- γ, TNF- α, and IFN- αwere detected according to the manufacturer’s instructions (Cell-Genebio, Hangzhou, China). EDTA anticoagulant blood vessels were centrifuged at 3000 rpm for 20 minutes. A low-speed centrifuge was used to collect the supernatant (a) for 200 g / 5 minutes, and we added microsphere buffer (H) at the same volume as the supernatant. A vortex mixer was used on the mixture which was then incubated in darkness for 30 minutes followed by vortex mixing. A 25 ul volume of the microsphere mixture was added to each experimental tube in order; 25 ul of the sample to be tested (plasma); fluorescent detection reagent (c); incubate in dark for 2.5 h at room temperature. Add 1 ml PBS solution, centrifugation at 200 g / 5 min; discard the supernatant; add 110 ul PBS solution to resuspend cells, flow cytometry detection. Fcap3.0 software analysis, five times within the batch repeated CV < 2%

#### Statistical analysis

The results of continuous measurements are presented as mean (SD) if they are normally distributed or median (IQR) if they are not normally distributed. Categorical variables are given as count (%). For laboratory results, we assessed whether the measurements were outside the normal range. We used SPSS (version 26.0) for all analyses.

### Patient and public involvement

This was a retrospective case series study and no patients were involved in the study design, setting the research questions, or the outcome measures directly. No patients were asked to advise on interpretation or writing up of results.

## Results

### 1. Clinical Characteristics and laboratory examination

All 11 COVID-19-infected critically ill pneumonia patients were admitted to the ICU. 10 (90.9%) of them were males. The median age was 58 years (IQR, 49-72 years). The median time from first symptoms to respiratory distress was 10 d (IQR, 7-13 days). All patients had fever, and the highest fever was 40°C, and the median fever temperature was 38.5°C (IQR, 38-39.6°C). Four (36.4%) patients had muscle soreness in the early stage of fever, and 9 (81.8%) had dry cough in the early stage. All patients had symptoms of hypoxia when they were admitted to ICU. Among them, 5(45.5%)patients had a history of Cardio-cerebrovascular disease, and 4(36.4%) patients had a history of diabetes (type II diabetes). All the patients had symptoms of low oxygen saturation and metabolic acidosis (Table 1). After entering the ICU, all the patients had at least one or more organ dysfunction or even failure manifestations. These included 9 patients with cardiac dysfunction, 8 with renal dysfunction, 6 with liver dysfunction and coagulation dysfunction, 5 with coagulant function abnormality, and 4 with multi-organ failure syndrome. 10 patients were treated with steroids and all patients had antiviral and anti-infective therapy (Table 2 and Table 3).

**Table 1:**
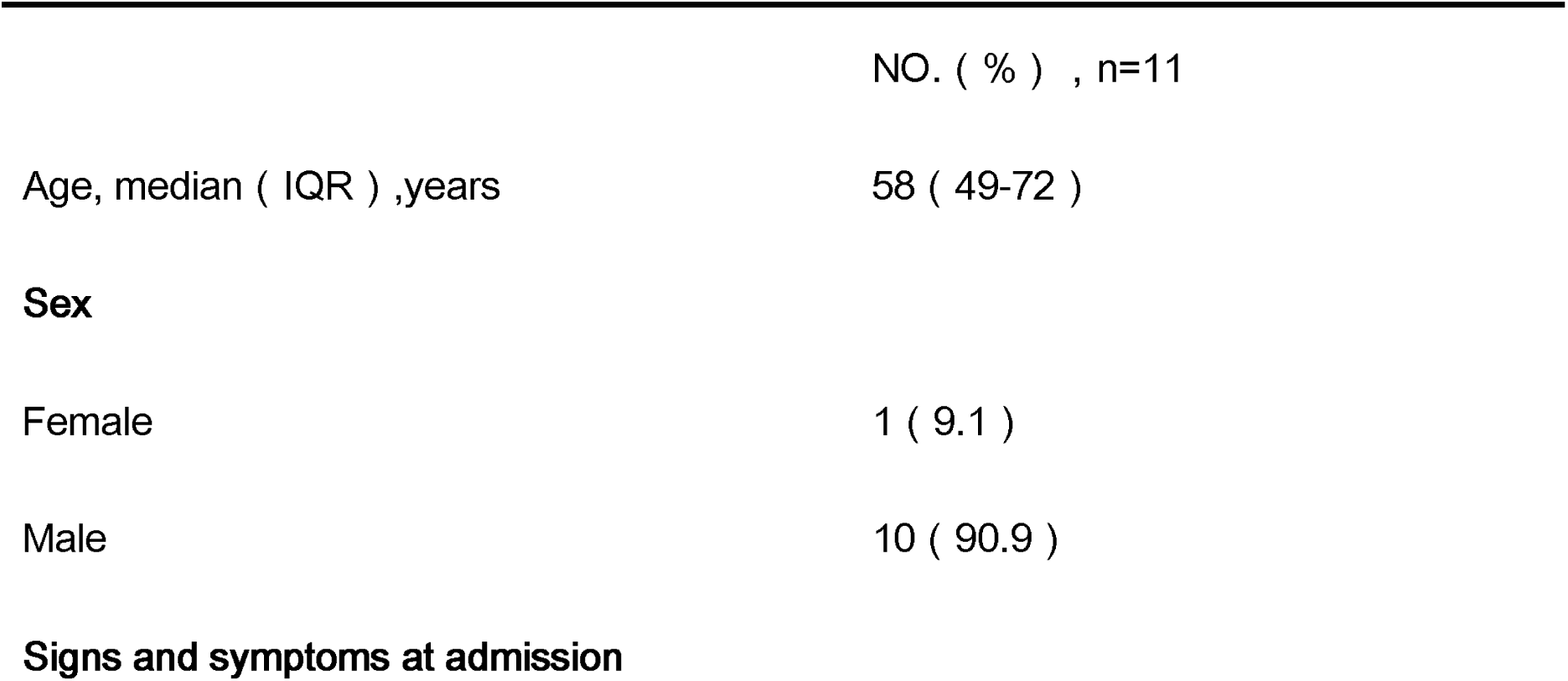
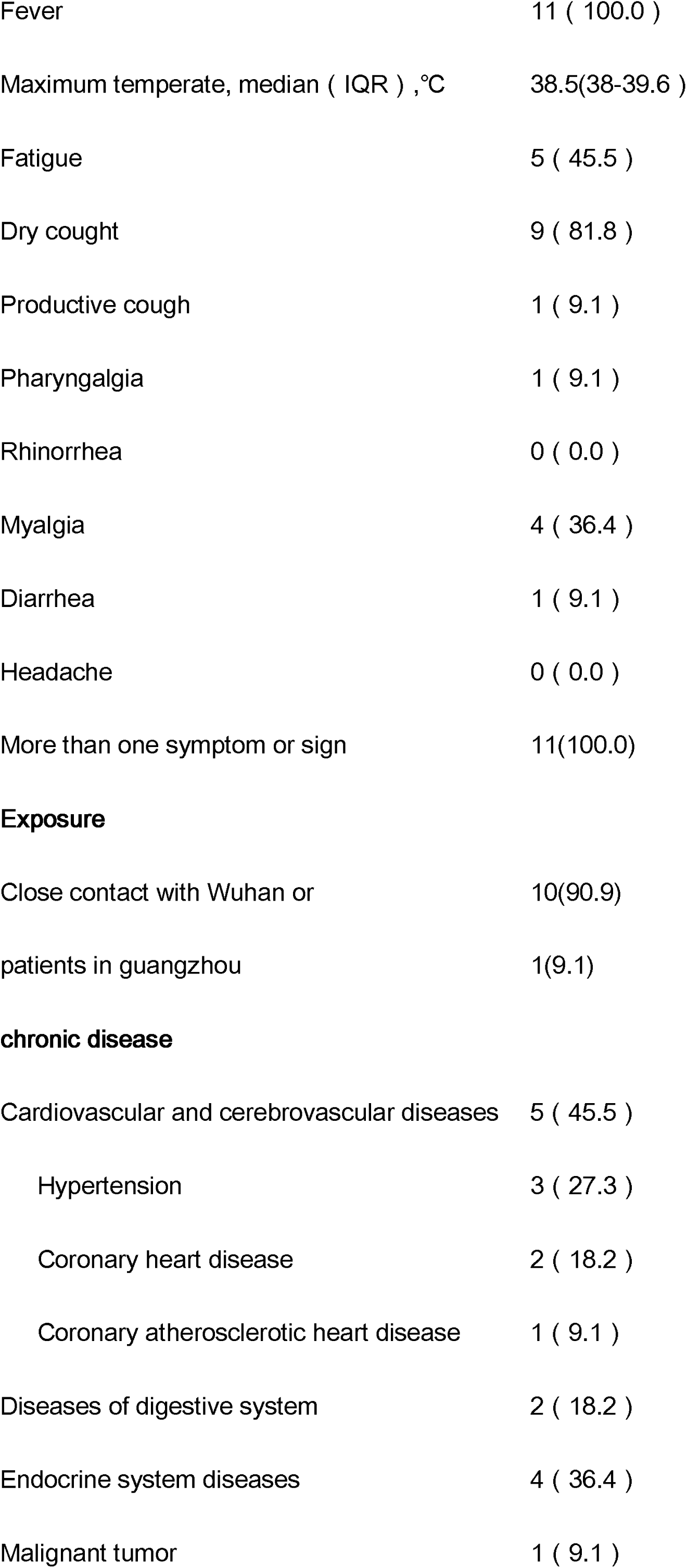
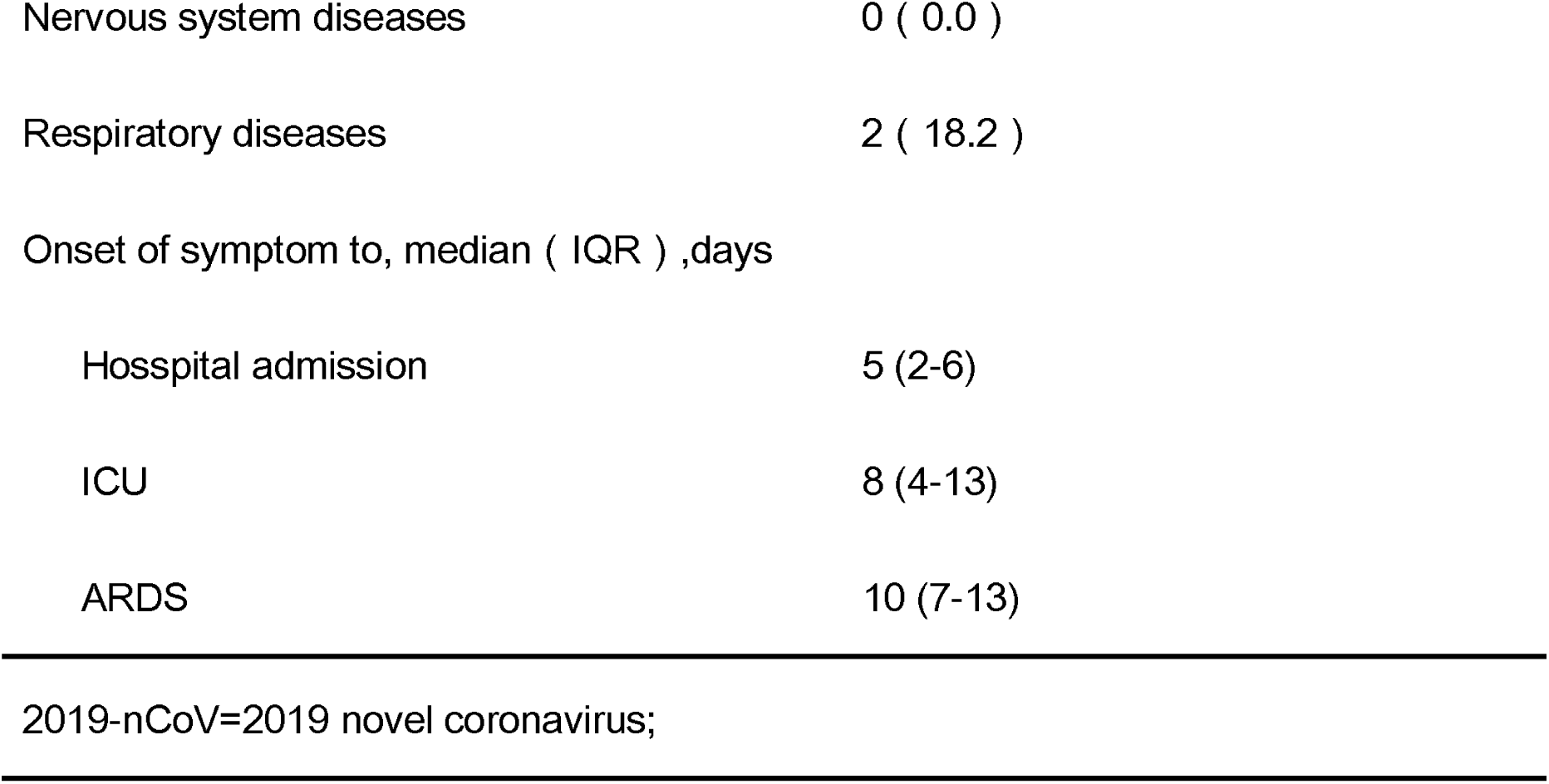
Demographics, baseline characteristics of COVID-19 Patients

|  | NO. ( % ) , n=11 |
| --- | --- |
| Age, median ( IQR ) ,years | 58 ( 49-72 ) |
| **Sex** |  |
| Female | 1 ( 9.1 ) |
| Male | 10 ( 90.9 ) |
| **Signs and symptoms at admission** |  |
| Fever | 11 ( 100.0 ) |
| Maximum temperate, median ( IQR ) , °C | 38.5(38-39.6 ) |
| Fatigue | 5 ( 45.5 ) |
| Dry cough | 9 ( 81.8 ) |
| Productive cough | 1 ( 9.1 ) |
| Pharyngalgia | 1 ( 9.1 ) |
| Rhinorrhea | 0 ( 0.0 ) |
| Myalgia | 4 ( 36.4 ) |
| Diarrhea | 1 ( 9.1 ) |
| Headache | 0 ( 0.0 ) |
| More than one symptom or sign | 11(100.0) |
| **Exposure** |  |
| Close contact with Wuhan or | 10(90.9) |
| patients in guangzhou | 1(9.1) |
| **chronic disease** |  |
| Cardiovascular and cerebrovascular diseases | 5 ( 45.5 ) |
| Hypertension | 3 ( 27.3 ) |
| Coronary heart disease | 2 ( 18.2 ) |
| Coronary atherosclerotic heart disease | 1 ( 9.1 ) |
| Diseases of digestive system | 2 ( 18.2 ) |
| Endocrine system diseases | 4 ( 36.4 ) |
| Malignant tumor | 1 ( 9.1 ) |
| Nervous system diseases | 0 ( 0.0 ) |
| Respiratory diseases | 2 ( 18.2 ) |
| Onset of symptom to, median ( IQR ) ,days |  |
| Hospital admission | 5 (2-6) |
| ICU | 8 (4-13) |
| ARDS | 10 (7-13) |

2019-nCoV=2019 novel coronavirus;

**Table 2:**
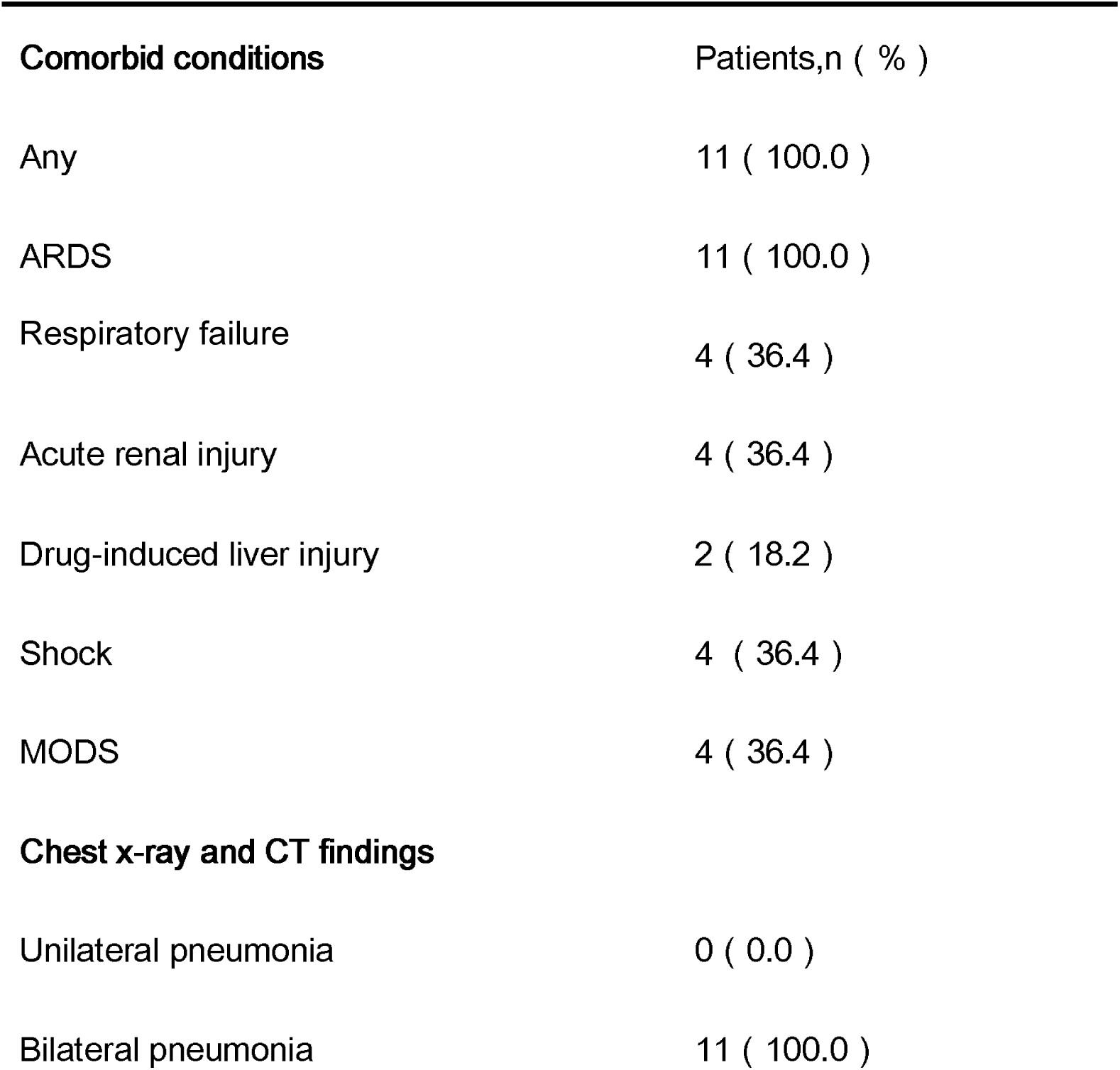
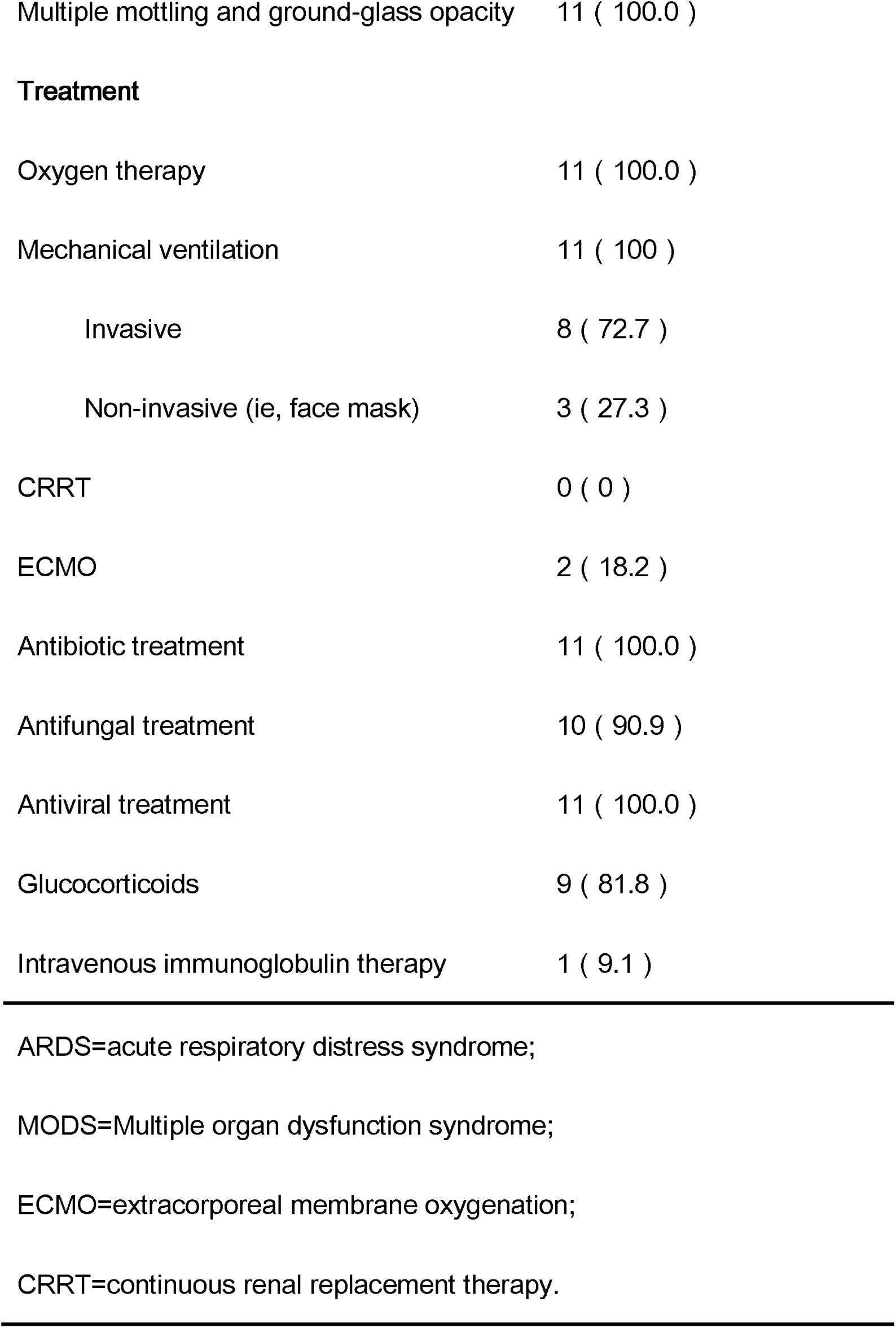
Clinical treatment of patients with COVID-19 pneumonia

**Table 3:** Laboratory results of patients with COVID-19 pneumonia

|  | Normal Rang | Median(IQR),<br>Patients (n=11) |
| --- | --- | --- |
| **Blood routine** |  |  |
| white blood cell count , $\times 10^9/L$ | 4.0-10.0 | 10.2 ( 6.5-12.4 ) |
| Neutrophil count , $\times 10^9/L$ | 1.8-8.0 | 9.6 ( 4.9-11.8 ) |
| Lymphocyte count , $\times 10^9/L$ | 0.9-5.2 | 0.3 ( 0.2-0.5 ) |
| Monocyte count , $\times 10^9/L$ | 0.16-1.0 | 0.4 ( 0.3-0.5 ) |
| Platelet count , $\times 10^9/L$ | 100-400 | 143 ( 113-210 ) |
| **coagulation function** |  |  |
| Prothrombin time, s | 11-14.5 | 15.1 ( 14.8-15.6 ) |
| Activated partial thromboplastin time, s | 28-42.8 | 37.6 ( 35.2-52.6 ) |
| D-dimer,(ng/ml FEU) | 68-494 | 1318 ( 673-4757 ) |
| **liver function** |  |  |
| Alanine aminotransferase , U/L | 5-40 | 24 ( 15.9-27.7 ) |
| Albumin, g/L | 35-55 | 33.6 ( 30.5-37.2 ) |
| total bilirubin , $\mu\text{mol/L}$ | 1.7-22.2 | 15.1 ( 11.2-20.4 ) |
| Direct bilirubin , $\mu\text{mol/L}$ | 0-6 | 4.9 ( 4.1-5.7 ) |
| **kidney function** |  |  |
| Serum glucose, mmol/L | 3.6-6.1 | 11.6 ( 7.72-16.03 ) |
| Serum urea nitrogen, mmol/L | 3.6-9.5 | 10.2 ( 7.4-17.3 ) |
| Serum creatinine, $\mu\text{mol/L}$ | 57.0–111.0 | 86.1 ( 62-146.6 ) |
| **myocardial injury markers** |  |  |
| Lactate dehydrogenase, U/L | 109-255 | 396.5 ( 357.6-529 ) |
| Creatine kinase, U/L | 10-190 | 154.5 ( 66.5-501.6 ) |
| Creatine kinase-MB, U/L | 3-25 | 13 ( 8-21 ) |
| Myoglobin, g/L | <70 | 62.1 ( 37.25-389.85 ) |
| Troponin I, µg/L | 0-0.04 | 0.01 ( 0-0.04 ) |
| Serum B-type natriuretic peptide precursor,pg/ml | <300 | 364.8 ( 147.2-746.9 ) |
| **infection related biomarkers** |  |  |
| Procalcitonin,ng/mL | 0.0-5.0 | 0.32 ( 0.095-0.435 ) |
| C-reactive protein ( CRP ) ,mg/dL | 0-0.6 | 12.06 ( 6.24-13.75 ) |
| **cytokines** |  |  |
| IL2,pg/ml | 0-5.71 | 1.57 ( 1.09-1.96 ) |
| IL4,pg/ml | 0-2.80 | 1.97 ( 1.64-2.12 ) |
| IL6,pg/ml | 0-5.30 | 26.87 ( 14.26-92.2 ) |
| IL10,pg/ml | 0-4.91 | 4.75 ( 3.83-11.19 ) |
| TNF-a,pg/ml | 0-2.31 | 1.7 ( 1.27-2.07 ) |
| IFN-γ,pg/ml | 0-7.42 | 1.46 ( 1.24-1.98 ) |
| **Immunocyte detection and absolute count** |  |  |
| CD3+CD45+ , % | 50-84 | 66 ( 59.8-68.4 ) |
| CD3+CD4+ , % | 26-61 | 44.1 ( 32.0-53.3 ) |
| CD3+CD8+ , % | 15-44 | 18.3 ( 6.9-32.8 ) |
| CD4+/CD8+ | 1.4-2.0 | 2.09 ( 1.05-7.7 ) |
| B cell ( CD3-CD19+ ) , % | 5-18 | 28.2 ( 11.8-30.3 ) |
| NKcell (CD3-CD16+CD56+) , % | 7-40 | 9.7 ( 3.7-12.4 ) |
| NKTcell (CD3+CD16+CD56+) , % | 2-13 | 2.8 ( 0.7-5.3 ) |
| Treg cell ( CD4+CD27+127- ) , % | 3-8 | 5.1 ( 3.3-7.8 ) |
| CD3+CD45+,absolute count, cells/μl | 955-2860 | 296 ( 169-355 ) |
| CD3+CD4+, absolute count, cell/μl | 550-1440 | 220 ( 101-308 ) |
| CD3+CD8+ absolute count , cells/μl | 320-1250 | 56 ( 33-141 ) |
| B cell ( CD3-CD19+ ) absolute count,<br>cells/μl | 90-560 | 81 ( 61-146 ) |
| NK cell (CD3-CD16+CD56+) absolute<br>count, cells/μl | 150-1100 | 34 ( 8-72 ) |
| NKTcell (CD3+CD16+CD56+) absolute<br>count, cells/μl | 40-300 | 10 ( 3-16 ) |

### 2. Immunological parameters analysis

Flow cytometry was used to detect the peripheral blood monocytes, including: CD3 (CD3 + CD45+), CD4 (CD3 + CD4+), CD8 (CD3 + CD8+), NK (CD3-CD16 + CD56 +), Tregs (CD4 + CD25 + CD127 low), and b-lymphocyte (CD3-CD45 +). In the first day being admitted to the ICU, the absolute counts of CD3, CD4, CD8, and NK in the peripheral blood were lower than the normal value: CD3 (IQR,169-355; range, 50–635 cells/μL), CD4 (IQR,101–308; range, 27–350 cells/μL), CD8 (IQR, 33-141; range, 21–277 cells/μL), NK immune cells (IQR,8-72; range, 5-170 cells/μl). Eight patients had a significant increase of the CD4/CD8 ratio (IQR, 1.05-7.7), which is an important index in autoimmune diseases (systemic lupus erythematosus, type I diabetes, and rheumatoid arthritis) and transplant rejection (16) (Table 3). Three had a decrease in B-lymphocytes, and two patients had a slight increase in Tregs (sFigure 1). After treatment, the CD4 in two patients and CD8 in three patients were increase to normal levels. In addition, IL-2, IL-4, IL-6, IL-10, TNF - α, and IFN - γ were detected in all of the patients. IL-6 was significantly increased in all of the patients (IQR, 14.26-99.2; range, 4.58–1182.91ng/L) (Table 3). The level of IL-10 was increased in five patients, while IL-4 and IFN - γ were increased in three and two patients, respectively. After treatment, IL-6 in three patients, IL-10 in four patients and IL-4, IFN - γ in all patients were returned to normal levels (sFigure 1).

### 3. Identification of CRSL in COVID-19-Infected Critically ill pneumonia patients

The main characteristics of CRS include a decrease of T cells and NK cells, an increase of IL-6, fever, organ and tissue dysfunction, and an abnormal coagulation function (20). All of the 11 ICU patients had pulmonary infection and fever. All had a significant decrease of circulating T cells, NK cells, and an increase of IL-6 secretion in peripheral blood. In addition, all the patients had one or more organ dysfunctions or failure, and seven patients had coagulation dysfunction. Eight patients showed a significant increase in the ratio of CD4/CD8, suggesting the possibility of an autoimmune response (Table 3 and Table 4). None of the patients had a known prior history of immune diseases. IL-6 level in three of these patients recovered to normal after infection treatment, and CT results shown the pulmonary shadows were disappeared. At the meantime, this three patients without evidence of extrapulmonary organ injury, and blood coagulation dysfunction, and two of them have been transferred out of ICU. There were eight of the 11 patients showed sufficient evidence in CRS features (Table 3 and Figure 1). After treatment, CD4, CD8, and NK cells were all recovered. In addition, IL-6 and IFN - γ were also decreased. At the same time, the organ function of the patients improved (Table 3 and sFigure 1).

**Table 4:**
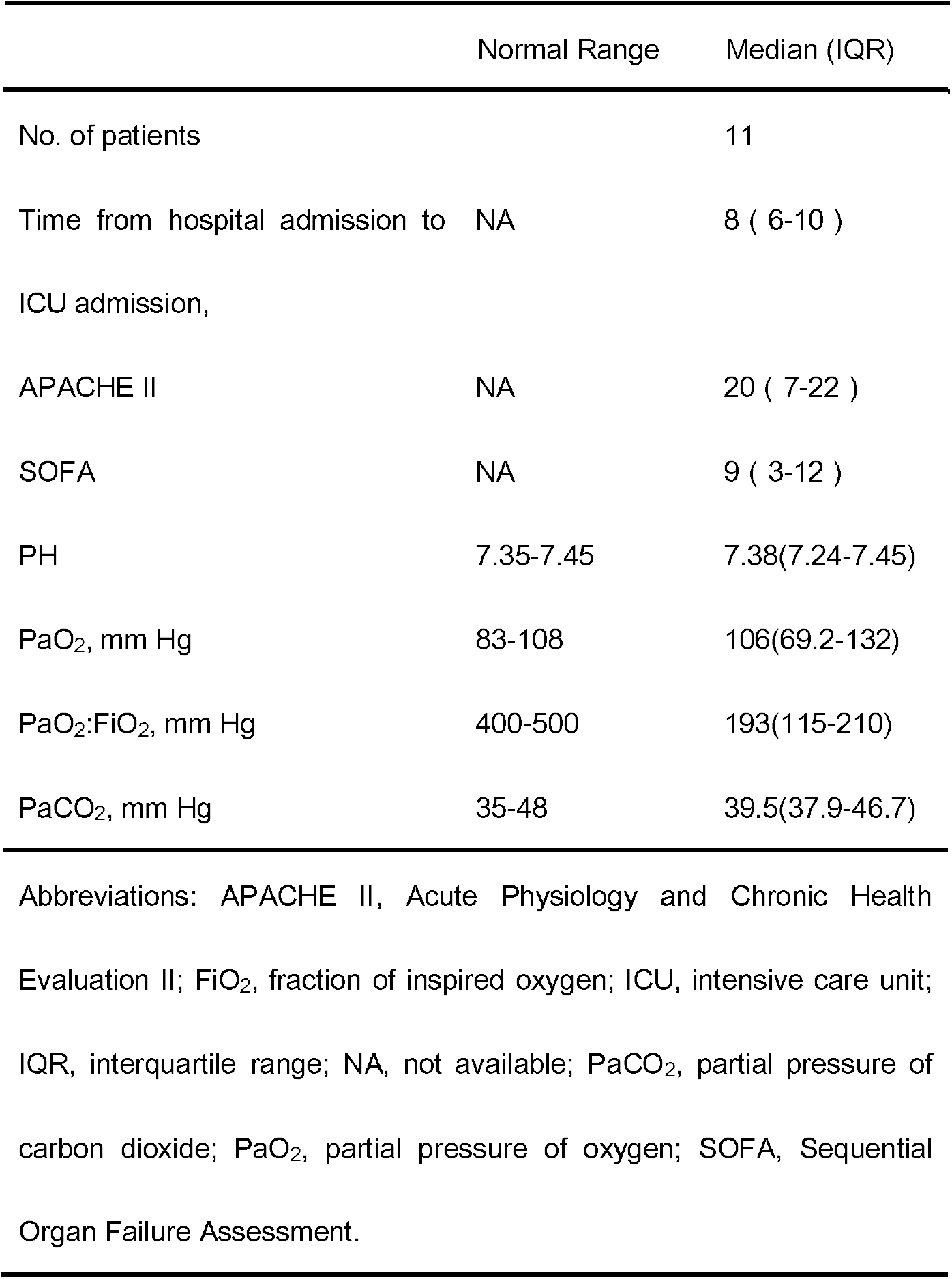
Severity of Illness Scores and Blood Gas Analysis of Patients Infected With 2019-nCoV pneumonia

| Normal Range | Median (IQR) |
| --- | --- |

|  |  |  |
| --- | --- | --- |
| No. of patients |  | 11 |
| Time from hospital admission to ICU admission, | NA | 8 ( 6-10 ) |
| APACHE II | NA | 20 ( 7-22 ) |
| SOFA | NA | 9 ( 3-12 ) |
| PH | 7.35-7.45 | 7.38(7.24-7.45) |
| PaO<sub>2</sub>, mm Hg | 83-108 | 106(69.2-132) |
| PaO<sub>2</sub>:FiO<sub>2</sub>, mm Hg | 400-500 | 193(115-210) |
| PaCO<sub>2</sub>, mm Hg | 35-48 | 39.5(37.9-46.7) |

Abbreviations: APACHE II, Acute Physiology and Chronic Health Evaluation II; FiO<sub>2</sub>, fraction of inspired oxygen; ICU, intensive care unit; IQR, interquartile range; NA, not available; PaCO<sub>2</sub>, partial pressure of carbon dioxide; PaO<sub>2</sub>, partial pressure of oxygen; SOFA, Sequential Organ Failure Assessment.

**Figure 1.**
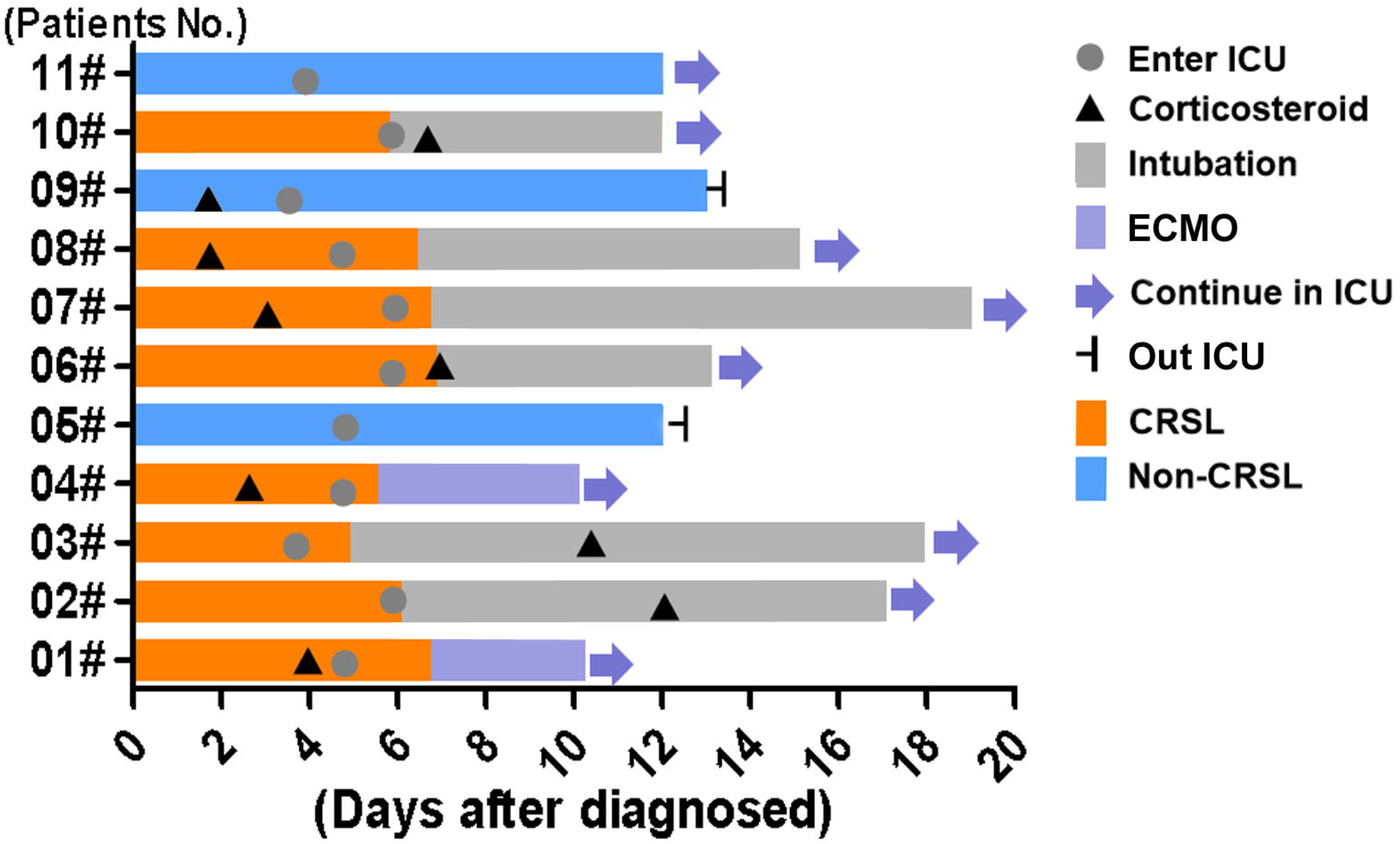
Timeline of disease course according to days from definite diagnosis of illness and days from Enter ICU. Before entering the ICU, nine people received corticosteroid therapy. In the treatment and nursing process of ICU, six patients were intubated, two patients were ECMO, and eight patients had CRS-like symptoms, two patients who had non-CRS-like symptoms were transferred out of ICU.

### 3. IL-6 was a sensitive indicator for the outcome of COVID-19 pneumonia with CRSL

All 11 patients were admitted to the ICU because of fever and ARDS. Among them, three patients received noninvasive ventilation, eight patients received invasive ventilation (including two ECMO treatment). Four people used medicine to cool down, 7 cases used physical hypothermy (ice compress). All of the patients were COVID-19 positive and received antiviral or anti-infective treatment, while nine received hormone treatment. During the treatment period, 9 patients had peripheral CD3, 9 patients had CD4, 8 patients had CD8, and 8 patients had NK cells increased, while seven patients had a decrease in CD3 / CD8 (Table 4 and Figure 2A). However, there were 3 patients (patient 02#, 03# and 07#) who with stable condition after ICU at the beginning, the IL-6 levels were rapid increase (increased 7.2 to 9.2 times) in one or two days(Figure 2B); the absolution number of CD4 and CD8 were decrease one day later than the increase of IL-6 in peripheral blood (Figure 2C and 2D). The PaO_2_ and PaO_2_/FiO_2_ were decrease within the next day after the rose of IL-6 (Figure 2E and 2F). At the meantime, the bedside chest radiograph results shown the disease progression (Figure 2G). This cases indicated IL-6 was a early indicators of CRSL in COVID-19-infected pneumonia.

**Figure 2.**
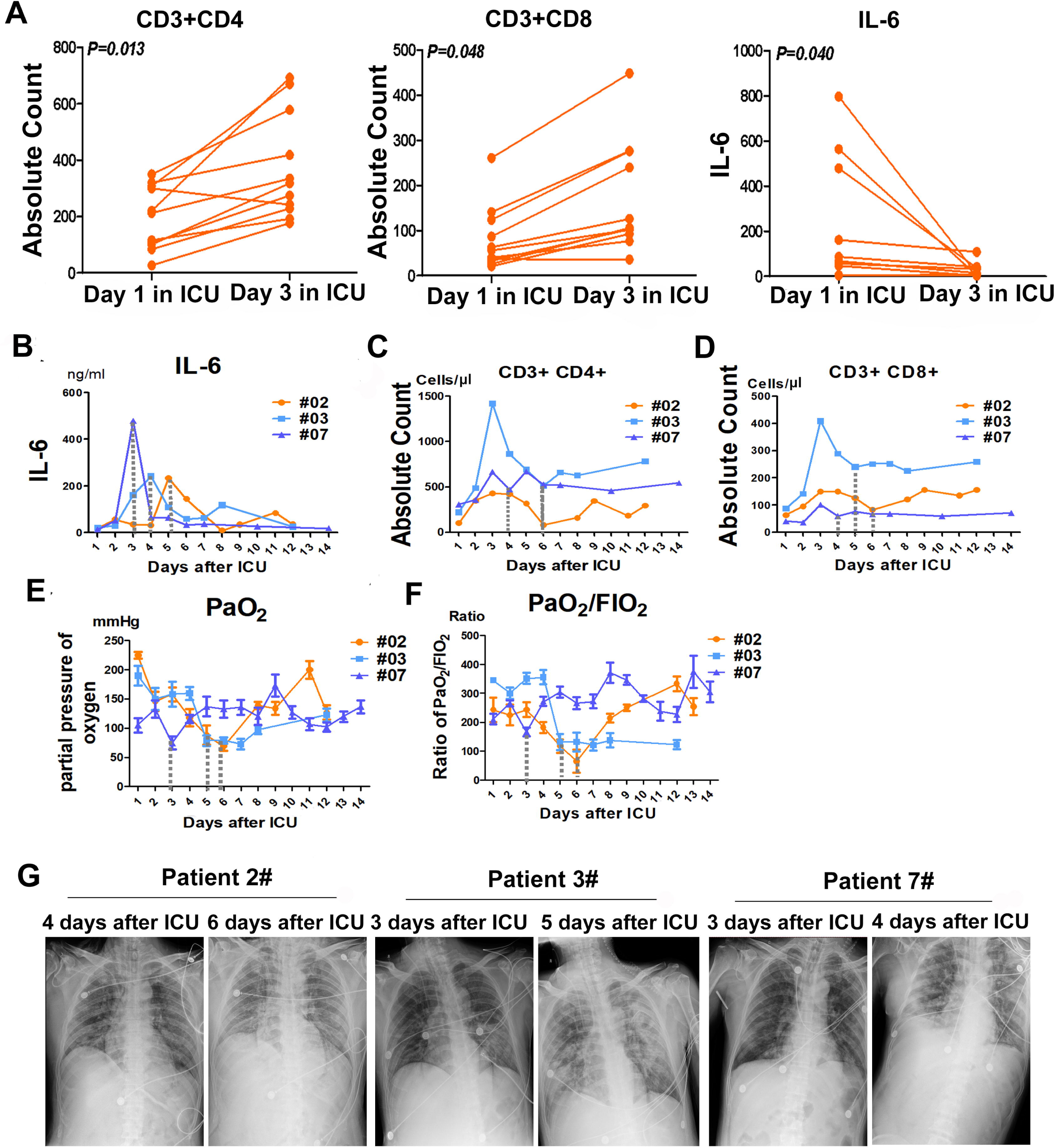
The relationship of Immunologic detection and CRSL after COVID-19 infected. A. The effect of effective treatment in ICU on immune cell subsets and cytokine IL6 Changes of immune cell subsets and cytokine IL-6 after effective treatment in ICU. The absolute counts of CD4+(P=0.013) and CD8+(P=0.048) cells were increased, while the inflammatory cytokines IL-6 (P=0.040) was decreased. B-F. Dynamic analysis of IL-6 levels,CD3+CD4+, CD3+CD8+ absolute counts, PaO_2_, and PaO_2_/FiO_2_ in 3 patients with severe pneumonia of covid-19.The absolute counts of CD3+CD4+ in patient#3, patient#2 and patient#7 at different dates during ICU (B-D). PaO_2_ and the oxygenation index PaO_2_ / FiO_2_have been in a fluctuating level in different dates during ICU (E and F). G. Chest x-rays of three COVID-19-infected pneumonia patients at indicated date.

### 4. The relationship of pulmonary inflammation and CRSL characteristics after COVID-19 infected

Serious pulmonary inflammation is a common symptom in critically ill COVID-19-infected pneumonia patients [13]. The large area of infection and inflammatory reaction will cause cytokine release syndrome (CRS)[14]. In our study, according to the patchy shadows or ground glass opacity distribution in the chest CT images, we were divided the patients into < 50% and ≥ 50% groups (Figure 3A). The numbers of CD4, CD8, and NK cells in the peripheral blood of were higher in the ≥ 50% groups than in ≥ 50% groups patients (Figure 3B, 3C and 3D). At the meantime, the level of IL-6 in peripheral blood was lower in the patients with the area of inflammatory ≥50 than in <50% in CT imaging (Figure 3E).

**Figure 3.**
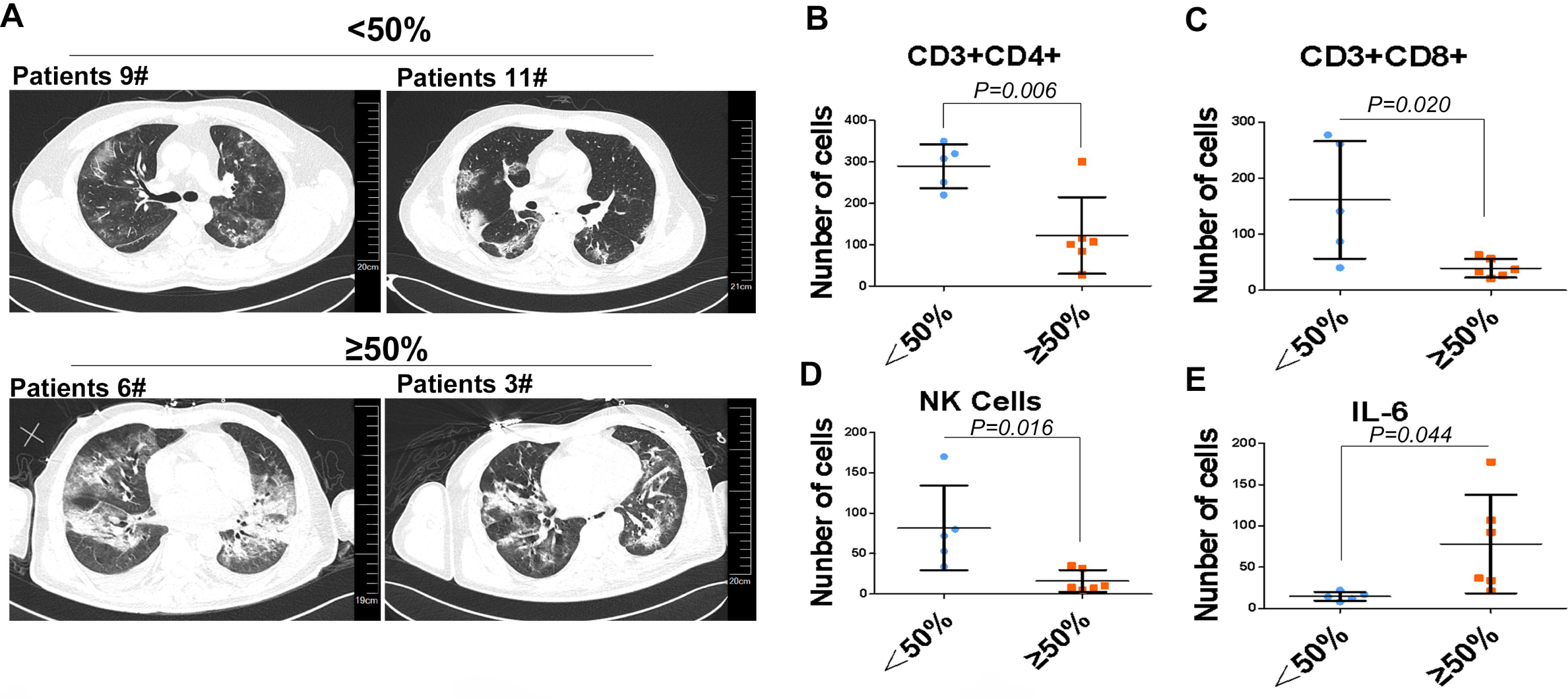
The relationship of pulmonary inflammation and CRSL characteristics after COVID-19 infected. A. Computed tomography of the chest of patients with COVID-19. According to the patchy or ground glass shadows in the chest CT images, the patients were divided into < 50% and ≥ 50% groups. B-E. The relationship between the shadow area of lung and the absolute count of CD3+CD4+, CD3 + CD8+, NK cells and the level of IL6 in plasma were compared. When the shadow area is less than 50%, the absolute counts of CD4 +(P=0.006), CD8 +(P=0.020), NK cells(P=0.016) is higher than the shadow area greater or equal to 50%. But the level of IL-6 (P=0.004) was in contrat.

## Discussion

This is a single-center retrospective study of 11 critically ill COVID-19 patients admitted to the ICU with persistent fever and ARDS. In addition to routine laboratory tests in the ICU, the number of immune cell subsets and cytokine levels in peripheral blood were analyzed in all patients. After analysis of the detection results, clinical characteristics, and diagnosis of all patients, we identified eight (72.7%) COVID-19-infected critical type pneumonia patients that had the clinical characteristics of CRSL.

Lung injury has a potential risk of CRS [17]. Cytokine release is common in immunotherapy, especially in cellular immunotherapy. The reasons for the occurrence of CRS are unclear. The main mechanism is that inflammatory cells, such as effector T cells and macrophages, accumulate rapidly from peripheral blood by chemokines and release a large number of cytokines into the blood when they kill tumor cells, viruses, or bacteria[18]. The disease is characterized by a marked increase in characteristic cytokines (such as IL-6), persistent fever, and organ and tissue damage[19]. The rapid chain reaction caused by cytokine release syndrome usually causes rapid organ immune related injury and acute functional failure[20].

We analyzed the clinical and immunological characteristics of 11 COVID-19-infected patients with severe pneumonia. We found that: Patients with severe pneumonia have different degrees of fever; peripheral blood CD4, CD8, NK, and other immune T cells decreased; CD4 / CD8 significantly increased, IL-6 significantly increased; multiple organ injuries were present; and coagulation dysfunction occurred. The concentration of IL-6 and the number of CD4, CD8, NK immune cells in the peripheral blood can be significantly reduced by improving ventilation, lowering body temperature, using anti-inflammatory treatment and other supportive treatment. The function of organs and tissues will be improved, which shows the importance of cytokines. COVID-19-infected patients with severe pneumonia have symptoms and manifestations similar to CRS. Therefore, we define this phenomenon as COVID-19 infection-related CRSL with the characteristics include: ① decrease of circulating blood CD4, CD8 T cells and NK cells; ② substantial increase of IL-6 cytokine in peripheral blood; ③ continuous fever; ④ organ and tissue damage caused by the cytokine related immune reaction, and the coagulation dysfunction. We also found that 8 of 11 (72.7%) critically ill patients in our study had characteristics consistent with cytokine-like release syndrome.

The concentration of IL-6 and the number of CD4, CD8 and NK immune cells in the peripheral blood can be reduced by improving ventilation, lowering body temperature, anti-inflammatory treatment, and other supportive treatment after ICU. The function of organs and tissues will be improved, which shows the dependence of cytokines. COVID-19-infected patients with severe pneumonia have evidence and manifestations similar to CRS. Large-scale lung injury caused by viral pneumonia is the inducing factor of COVID-19-infected CRS. Because large-scale lung injury is a typical feature of critical patients, COVID-19-infected CRS may be a common phenomenon in critically ill patients. We found that a large area of lung injury (≥50%) with an decrease of CD4, CD8 (Lower than 50% minimum normal range) and increase of IL-6 in peripheral blood was the highest risk factor of CRSL. After the COVID-19-infected pneumonia in the critically ill patients, the change of IL-6 levels were one or two days earlier than the absolution number of CD4 and CD8 decreased in peripheral blood. This indicated IL-6 may works as an early indicator of CRSL in COVID-19-infected pneumonia.

This study has several limitations. First, we only studied 11 critical patients in a single center and we were unable to conduct a group comparison study. Additional data from critical patients in China would be useful. Second, due to the strong infectivity of the virus, we were not able to construct a COVID-19-infected pneumonia animal model. A validated model could help clarify the molecular mechanism of CRS caused by viral pneumonia. Third, we have not finished the clinical experiment on the treatment of COVID-19-infected pneumonia with IL-6 monoclonal antibody.

## Conclusions

We identified and defined the CRSL in 11 critically ill patients with COVID-19-infected pneumonia. We found that a large area of lung injury by COVID-19-infected was often accompanied by the increase of IL-6 and decrease of CD4, CD8 and NK immune T cells in peripheral blood and was the risk factor of CRS. IL-6 was an early indicator of CRSL in COVID-19-infected pneumonia. We also found that reduce injury to the lung is a useful method to prevent and improve COVID-19-infected pneumonia-related CRSL in critically ill pneumonia patients.

## Data Availability

a.) Basic information of the patient: name, gender, age, hospitalization number, diagnosis, date of admission, condition, laboratory examination results, chest CT scan, nursing care, and treatments (i.e. antiviral treatment, corticosteroid treatment, and respiratory support. The records were taken by a team of trained doctors, and the data were obtained from electronic medical records.
b. Data generated by bedside monitor: heart rate, respiration, invasive / non-invasive blood pressure, blood oxygen saturation, hemodynamic monitoring.
c. Medical order execution information: administration times for all medicines, drug name, dose, concentration, route, automatically generated in and out quantity. These data were automatically collected by the ICU clinical information system.

## Patient and Public Involvement

For Research articles only

## Footnotes

### Contributors

wang-wj,wu-sp and liu-xq contributed equally to this article. liu-xq conceptualised the paper. wang-wj and wu-sp analysed the data, with input from lie-py, h-ly, chen-sb, nong-lb, cheng-ll and lin-yp. wang-wj wrote the initial draft with all authors providing critical feedback and edits to subsequent revisions. All authors approved the final draft of the manuscript. he-jx is the guarantor. The corresponding author attests that all listed authors meet authorship criteria and that no others meeting the criteria have been omitted.

### Ethical approval

This study was approved by the Ethics committee of the First Affiliated Hospital of Guangzhou Medical University

### Patient consent

Obtained.

### Data sharing

No additional data available.

### Transparency

The lead authors and manuscript’s guarantor affirm that the manuscript is an honest, accurate, and transparent account of the study being reported; that no important aspects of the study have been omitted; and that any discrepancies from the study as planned have been explained.

### Dissemination to participants and related patient and public communities

No study participants were involved in the preparation of this article. The results of the article will be summarised in media press releases from the Guangzhou Medical University and presented at relevant conferences.

sFigure 1. A-E. Dynamic analysis of absolute count of immune cell subsets in 11 patients with severe pneumonia of covid-19.

F-J. Dynamic analysis of serum inflammatory cytokines in 11 patients with severe pneumonia of covid-19.

## REFERENCES

1. Huang C, Wang Y, Li X, et al. Clinical features of patients infected with 2019 novel coronavirus in Wuhan, China. Lancet 2020;395: 497–506.

2. Chan JF, Yuan S, Kok KH, et al. A familial cluster of pneumonia associated with the 2019 novel coronavirus indicating person-to-person transmission: a study of a family cluster. Lancet 2020;395: 514–23.

3. Wu F, Zhao S, Yu B, et al. A new coronavirus associated with human respiratory disease in China. Nature 2020; published online Feb 3. DOI:10.1038/s41586-020-2008-3.

4. Li Q, Guan XH, Wu P, et al. Early Transmission Dynamics in Wuhan, China, of Novel Coronavirus–Infected Pneumonia. N Engl J Med. 2020 Jan 29.

5. Tan WJ, Zhao X, Ma XJ, et al. A novel coronavirus genome identified in a cluster of pneumonia cases — Wuhan, China 2019–2020. China CDC Weekly 2020; 2: 61–2.

6. National Health Commission of the People’S Republic Of China. http://www.nhc.gov.cn/xcs/yqfkdt/202002/4a1b1ec6c03548099de1c3aa935d04fd.shtml. 2020.

7. Interpretation of “Guidelines for the Diagnosis and Treatment of Novel Coronavirus (2019-nCoV) Infection by the National Health Commission (Trial Version 5)”. February 2020 Zhonghua yi xue za zhi 100:E001.

8. Butt Y, Kurdowska A, Allen TC. Acute Lung Injury: A Clinical and Molecular Review. Arch Pathol Lab Med. 2016 Apr;140(4):345–50.

9. Mokra D, Kosutova P. Biomarkers in acute lung injury. Respir Physiol Neurobiol. 2015 Apr;209:52–8.

10. Paolone S. Extracorporeal Membrane Oxygenation (ECMO) for Lung Injury in Severe Acute Respiratory Distress Syndrome (ARDS): Review of the Literature. Clin Nurs Res. 2017 Dec;26(6):747–762.

11. Wang D, Hu B, Hu C, et al. Clinical Characteristics of 138 Hospitalized Patients With 2019 Novel Coronavirus-Infected Pneumonia in Wuhan, China. JAMA. 2020 Feb 7.

12. Panitchote A, Mehkri O, Hastings A, et al. Factors associated with acute kidney injury in acute respiratory distress syndrome. Ann Intensive Care. 2019 Jul 1;9(1):74.

13. Xu X, Yu C, Zhang L, Luo L, Liu J. Imaging features of 2019 novel coronavirus pneumonia. Eur J Nucl Med Mol Imaging. 2020 Feb 14.

14. Zhao H, Chen H, Xiaoyin M, et al. Autophagy Activation Improves Lung Injury and Inflammation in Sepsis. Inflammation. 2019 Apr;42(2):426–439.

15. Tanaka T, Narazaki M, Kishimoto T. Immunotherapeutic implications of IL-6 blockade for cytokine storm. Immunotherapy. 2016 Jul;8(8):959–70.

16. Carvajal Alegria G, Gazeau P, Hillion S, et al. Could Lymphocyte Profiling be Useful to Diagnose Systemic Autoimmune Diseases? Clin Rev Allergy Immunol. 2017 Oct;53(2):219–236.

17. Kogure Y, Ishii Y, Oki M. Cytokine Release Syndrome with Pseudoprogression in a Patient with Advanced Non-Small Cell Lung Cancer Treated with Pembrolizumab. J Thorac Oncol. 2019 Mar;14(3):e55–e57.

18. Shimabukuro-Vornhagen A, Gödel P, Subklewe M, et al. Cytokine release syndrome. J Immunother Cancer. 2018 Jun 15;6(1):56.

19. Uciechowski P, Dempke WCM. Interleukin-6: A Masterplayer in the Cytokine Network. Oncology. 2020 Jan 20:1–7.

20. Liu E, Marin D, Banerjee P, et al. Use of CAR-Transduced Natural Killer Cells in CD19-Positive Lymphoid Tumors. N Engl J Med. 2020 Feb 6;382(6):545–553.

